# Implementing a program to improve handoffs and reduce adverse events in Paediatric Intensive Care Units in Argentina: a stepped wedge trial

**DOI:** 10.1101/2020.09.07.20188672

**Authors:** Jorro-Barón Facundo, Suarez-Anzorena Inés, Burgos-Pratx Rodrigo, Demaio Noelia, Penazzi Matias, Rodriguez Gisela, Rodriguez Ana Paula, Velardez Daniel, Ábalos Silvina, Lardone Silvina, Olivieri Joaquín, Gallagher Rocío, Rodriguez Rocio, Gibbons Luz, Landry Luis Martin, Garcia-Elorrio Ezequiel

## Abstract

**Introduction:** An effective and standardized communication anticipates and limits the appearance of possible adverse events.

**Objective:** To evaluate the effect of the implementation of a handoff program in reducing the frequency of adverse events (AE) in Paediatric Intensive Care Units (PICUs).

**Methods:** Facility-based, cluster randomised stepped wedge trial in six Argentine PICUs, with more than 20 admissions per month. The intervention comprised a Spanish version on the I-PASS Handoff Bundle consisting of a written and verbal handoff using mnemonics, an introductory workshop with teamwork training, an advertising campaign, simulation exercises and observation and standardized feedback of handoffs.

**Results:** We recruited 6 cluster PICUs in 5 hospitals. We reviewed 1465 medical records (MR). We did not observed differences in the rates of preventable AE per 1000 days of hospitalization (control 60.4 [37.5 - 97.4] vs. intervention 60.4 [33.2 - 109.9], *p* = 0.3568, RR:1.21 [CI95%:0.80 - 1.83]), and no changes in the categories or types of AE. We evaluated 847 handoffs. Compliance with all items in verbal and written handoff was significantly higher in the intervention group. We observed a longer time per patient to complete the handoff in the intervention group (7.29 minutes [5.77 - 8.81] vs. 5.96 [4.69 - 7.23]; p < 0.0002, RR:1.33 [CI95%:0.64 - 2.02]), without changes in the whole time used for handoff (control: 35.7 [29.6 - 41.8] vs. intervention: 34.7 [26.5 - 42.1]; p = 0.4900, RR:1.43 [CI95%:-2.63 - 5.49]). Perception of improved communication from provider didn’t show changes.

**Conclusions:** After the implementation of the I-PASS bundle, improvement in the quality of handoffs was observed. Nevertheless, no differences were observed in the frequency of AE, nor in the perception of improved communication.

## INTRODUCTION

In clinical settings, the effectiveness of communication is essential and must be considered as an interaction process(1). Communication errors represent the third cause of sentinel events(2), over half of which involve handoff failures(3). Handoff is defined as the exchange of information between health professionals about a patient, which is accompanied by a change in control or responsibility in the decisions of their care(4). It is estimated that a typical teaching hospital may experience more than 4,000 handoffs per day(5). The Joint Commission has established standardized transmission of information as a patient safety goal and advocates organizations to implement “a standardized approach to hand-off communications, including an opportunity to ask and respond to questions”(3).

Previous studies have shown that effective and standardized communication between caregivers in handoff is essential for patient safety, anticipates and limits possible errors(6–11). Different tools have been proposed as models to standardize the transmission of information during handoffs, many of them are acronyms to facilitate their use(12). The use of checklists was introduced to carry out handoffs between professionals in areas such as emergency rooms, wards, critical care units, and operating rooms. It was also shown that it not only contributes to reducing the length of handoffs, but also improves the quality of the information and the subsequent care of the patient(9). The implementation of an intervention including teamwork and a structured handoff has previously shown to decrease the number of adverse events (AE)(7).

The trigger tool methodology has proven useful in detecting AE. A tool called Global Assessment of Paediatric Patient Safety (GAPPS) has been described in the paediatric population, which proved to be reliable in measuring the incidence of AE(13). Trigger tools have shown that they can actively detect AE, through automated processes, at a higher rate than with the usual passive methods. Although GAPPS can be used to capture a wide variety of damage, these AEs are still discrete, easily quantifiable, and evident. However, children in the hospital setting are exposed to other more insidious harm. AEs produced by the delivery of unnecessary care may not be so easy to detect, because their effects may be subtle or may only be evident through long-term results (e.g. an unnecessary CT scan that triggers a leukaemia in the distant future (13,14).

There are only a few studies about handoffs quality and AE in low and middle-income countries, and none of them have rigorously evaluated the effectiveness of handoff-improvement programs. Stepped-wedge, cluster-randomised, controlled designs enabled both phased implementation and the use of established statistical approaches to compare control and intervention groups while minimizing the potential for bias and confounding(15).

We aimed to assess the effectiveness of a standardized handoff intervention to reduce the frequency of AE in Paediatric Intensive Care Units (PICUs) in a middle-income country.

## METHODS

### Study design

We conducted a facility-based, cluster randomized controlled trial with a stepped-wedge design in six PICUs between July 2018 and May 2019 (11 months). All participating units began as control practices without the trial intervention (three months). As the trial progressed, units were allocated randomly to receive the intervention in prespecified time periods (one month per step). This process continued until all participating clusters received the intervention. At the end of the study, all the clusters were exposed to the intervention for a period of at least 4 months.

### Randomisation

The unit of randomisation was the PICU. Sites were assigned to one of five start dates by the study statistician via a computer-generated list of random numbers. Concealment of the intervention starting date was maintained up to fifteen days previous to launch the intervention at each PICU, because the preparatory activities were needed it. The flowchart of the allocated sequence and period is described in Supplementary Appendix 1.

### Participants

The study was conducted in six PICUs of five public hospitals in three provinces of Argentina. Eligibility criteria for PICUs were the absence of a standardized handoff program in place and having at least 20 admissions per month.

### Intervention

A formative research was conducted to identify barriers and facilitators for the implementation of the I-PASS handoff program in order to calibrate the intervention in each participant sites. Information was gathered from 17 senior level health care professionals from the participant PICUs through in-depth interviews.

For this study we implemented the Spanish I-PASS bundle used in previous studies in our setting(16,17), consisting of ten elements: 1) the I-PASS mnemonics (Illness severity, Patient summary, Action list, Situation awareness and contingency plans, Synthesis by the receiver) which served as an anchoring component for verbal and written information; 2) an introductory 2-hour workshop with key content about quality of handoffs and training in teamwork; 3) five tools of teamwork training of TeamSTEPPS program -an evidence-based program aimed at optimising performance among teams of health care professionals -: cross monitoring, brief, debrief, huddle and check-back) from(18); 4) a written handoff I-PASS based form; 5) a 1-hour simulation and role-playing emphasizing the elements of the workshop; 6) a faculty development program; 7) a self-learning module to reinforce the components of the mnemonics; 8) direct-observation tools used by the faculty to provide feedback to physicians; 9) and an advertising campaign with printed material, posters and stickers with the IPASS logo and mnemonics for process and culture change; 10) a standardized I-PASS format written handoff template (6,7,19,20).

Each site also maintained an implementation log that was reviewed regularly to ensure adherence to each component of the handoff program. Biweekly meetings were held with each PICU to reinforce implementation strategies according to the difficulties presented and to ensure adherence to each component of the handoff program.

### Measurement of study outcomes

Adverse events were the primary outcome. Two independent reviewers, following a structured methodology, assessed AE. Reviewers were trained in the GAPPS process. A primary reviewer (a PICU staff, not necessarily physician) evaluated the selected medical records (MR) using the GAPPS list of 37 possible manual triggers using the following order: a) evaluation of the discharge and progress notes, b) prescriptions, and c) nursing progress sheets. The triggers were clues that suggest a possible AE. When primary reviewers identify a trigger (e.g., naloxone administration), the MR was reviewed to determine whether an AE might had happened (e.g., hypopnea from opioid overdose resulting from a prescription error) or not (e.g., naloxone used to reverse a heroin overdose). The primary reviewers presented the suspicions of AE to the secondary reviewers (a PICU staff, physician), who independently evaluated whether an AE had occurred and its’ severity too. Following so, all of the reviewers reached a consensus in every AE with an initial disagreement (21). During the review of the MR, every AE without a prior trigger was identified and also reported in the study. Up to 30 MRs in each cluster per month were reviewed.

The secondary outcome was the compliance with items of adequate verbal and written handoff and was assessed by direct observation. All physicians were observed at least once per month presenting or receiving a handoff. The observer completed an evaluation form about the compliance with the elements of a good quality handoff (Supplementary Appendix III).

Physicians were surveyed about patient safety culture, emphasizing the communication dimension using Surveys on Patient Safety Culture (SOPS) from the Agency for Healthcare Research and Quality (ARHQ) translated and validated in Spanish, previous and after the intervention was delivered(22).

### Sample size

Sample size was estimated aiming to reduce the preventable AE (primary outcome) from 12% to 4.5%(17). Assuming an ICC of 0.01, a number of steps = 5, a cluster size per step = 40, a power = 90% and an alpha Level = 5%, so the total number of clusters needed was six.

### Data Management and Statistical Analysis

Data collectors were trained in GAPPS tool use and handoff observation, using specifically designed data forms.

Analyses were performed according to the intention-to-treat principle. To evaluate compliance with verbal and written IPASS, each PICU was used as the unit of analysis. The results collected from the observation forms were grouped into a dichotomous scale for statistical analysis considering adherence to quality elements described as: almost always, always; and non-adherence: sometimes, almost never, never. The percentage of each of the variables reported by the PICU was calculated according to the time of the intervention. The median and interquartile range (25% quartile - 75% quartile) were described in both control and intervention groups. The effect of the intervention was evaluated using the absolute difference of the median percentages and was tested using the Wilcoxon test corrected for continuity.

AE were calculated for every 1000 days of hospitalization. To estimate the effect of the intervention, a generalized mixed linear model was adjusted assuming a negative binomial distribution. The size effect was adjusted by time trend, including the variable of the month of the study as a fixed effect in the model. Estimated rates with their confidence interval were reported in the control group and in the intervention group. The measure of effect was estimated using the ratio of both rates (intervention / control).

For the analysis of the AHRQ Surveys on Patient Safety Culture, the Likert scale was dichotomized, and the items were interpreted as a positive answer to each question: agree and strongly agree; and as a negative answer: strongly disagree, disagree and neither agree nor disagree.

Descriptive statistics were used to report all of the variables of interest. The analysis was performed in R for Windows (The R Foundation).

## RESULTS

During the study period, we reviewed 1465 MR with a total of 15842 PICU / patient days: 767 in the control period and 698 in the intervention period, in 6 different clusters (Figure 1). The MRs reviewed in each cluster and period are shown in the Supplementary Appendix II. We also observed 847 handoffs that yielded 5260 unique patient handoffs for evaluation. Patients’ baseline characteristics were similar in the control and intervention groups (Table 1).

**Figure 1.**
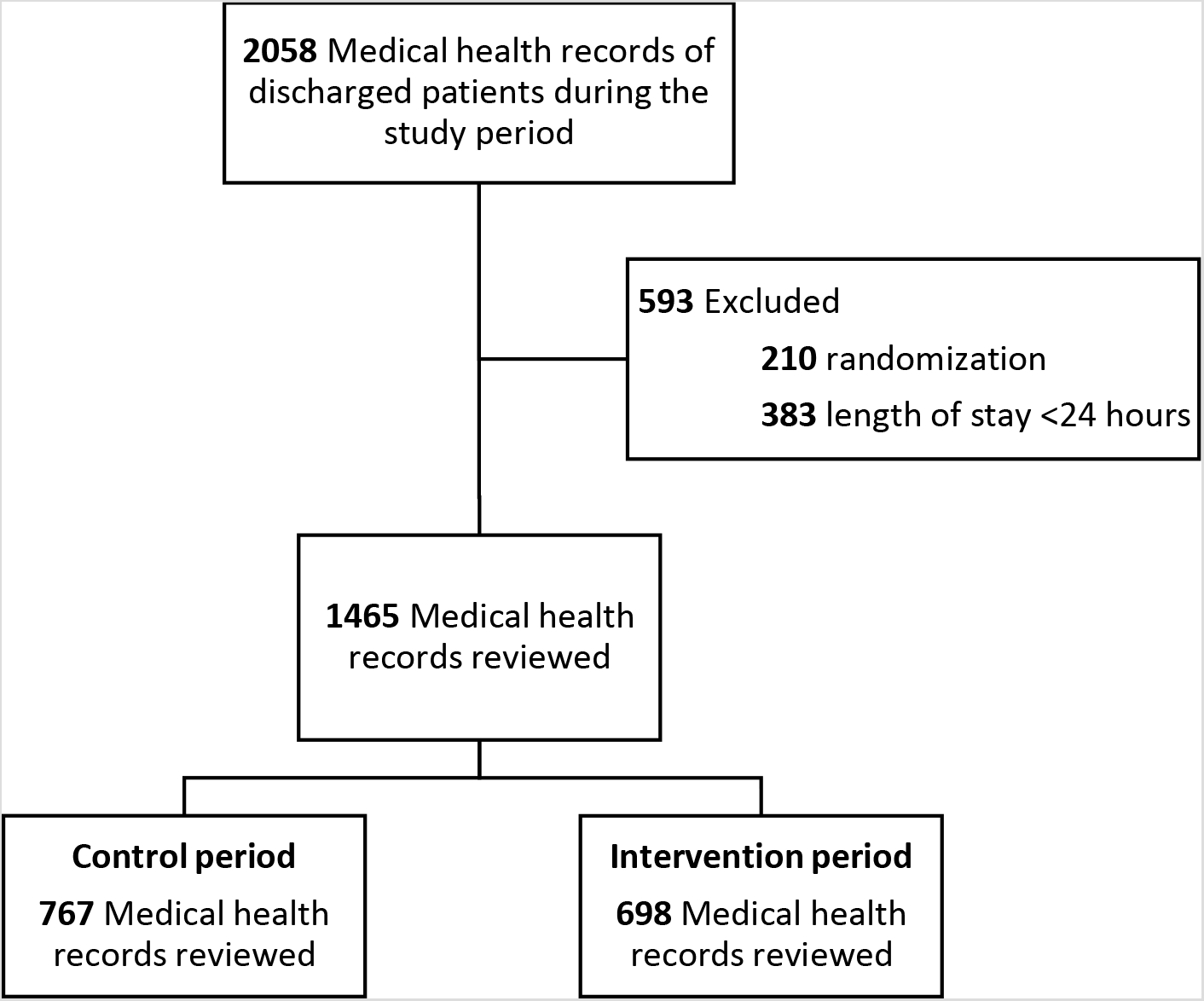
Consort diagram

**Table 1.** Patient baseline characteristics.

|  | Control<br>(N=767) | Intervention<br>(N=698) |
| --- | --- | --- |
| Age (months and SD) | 52.9±58.9 | 58.2±59.8 |
| Male sex (%) | 58.9 | 56.4 |
| Length of PICU stay (days and SD) | 10.7±16.8 | 10.9±21.2 |
| Paediatric Index of Mortality II score (mean and SD) | 5.5 ±10.6 | 5.9±12.3 |
| Mortality (%) | 14.5 | 17.1 |
| Destination on discharge - % (N) |  |  |
| Home | 4.0 (31) | 4.9 (34) |
| Ward | 76.1 (584) | 78.9 (551) |
| Rehab centre | 0.7 (5) | 0.4 (3) |
| Other institution | 3.7 (28) | 3.3 (23) |
| Homecare | 0.1 (1) | 1.0 (7) |
| Dead | 4.8 (37) | 5.7 (40) |
| Other | 9.7 (74) | 4.6 (32) |
| Not registered | 0.9 (7) | 1.2 (8) |

### Adverse events

The rate of preventable AE per 1000 days of hospitalization was similar in both arms, 60.4 [37.5; 97.4] in the control group and 60.4 [33.2; 109.9] in the intervention group, p = 0.3568, difference 1.21 [95% CI: 0.80; 1.83]. No differences were observed in AE per 1000 days of hospitalization, nor in the damage produced by the AE, or type of AE in control versus intervention periods (Table 2). AEs occurred as a result of hospital-acquired infections, care related (no medications or procedures), medication related, related to procedures and related to diagnosis, with no differences between study periods. The detail of each AE collected were in the Supplementary Appendix 2.

**Table 2.** Effect of the intervention on the primary and secondary outcomes (measured per 1000 days of hospitalization)

|  | Rates for 1000 days of hospitalization<br>(CI) |  | Rate of fees<br>(Intervention/Control)<br>(CI) | p-Value |
| --- | --- | --- | --- | --- |
|  | Control | Intervention |  |  |
| Total Adverse Events | 86.0<br>(60.0 - 123.3) | 93.7<br>(57.9 - 151.6) | 1.09<br>(0.84 - 1.42) | 0.5217 |
| Preventable Adverse Events | 60.4<br>(37.5 - 97.4) | 60.4<br>(33.2 - 109.9) | 1.0<br>(0.74 - 1.34) | 0.9982 |
| Preventable AE with temporary damage to the patient with intervention requirement | 18.8<br>(10.2 - 34.8) | 17.8<br>(7.8 - 41.0) | 0.95<br>(0.6 - 1.51) | 0.8200 |
| Preventable AE with temporary damage to the patient requiring admission to / prolongation of the hospitalization | 26.0<br>(16.1 - 42.0) | 20.2<br>(10.0 - 40.7) | 0.78<br>(0.51 - 1.18) | 0.2328 |
| Preventable AE with permanent damage to the patient | 1 event | 5 events | - | - |
| Preventable AE which required intervention to maintain life | 8.5<br>(2.8 - 26.2) | 16.0<br>(3.5 - 72.4) | 1.88<br>(0.82 - 4.31) | 0.1365 |
| Preventable AE with patient death | 3 events | 5 events | - | - |
| Medication related | 10.4<br>(4.5 - 24.0) | 10.8<br>(3.3 - 36.0) | 1.04<br>(0.52 - 2.09) | 0.9100 |
| Related to procedures | 9.7<br>(4.2 - 22.3) | 7.7<br>(2.1 - 28.0) | 0.79<br>(0.35 - 1.77) | 0.5689 |
| Care related (no medications or procedures) | 11.7<br>(5.2 - 26.0) | 9.9<br>(3.6 - 27.1) | 0.85<br>(0.52 - 1.39) | 0.5205 |
| Related to diagnosis | 1.6<br>(0.3 - 7.8) | 5.4<br>(0.6 - 48.5) | 3.39<br>(0.93 - 12.35) | 0.0643 |
| Hospital-acquired infections | 20.9<br>(14.4 - 30.3) | 19<br>(9.8 - 36.6) | 0.91<br>(0.58 - 1.41) | 0.6658 |
| Falls | 0 events | 0 events | - | - |
| Others | 0 events | 0 events | - | - |

### Assessment of written and oral handoffs

The adherence to an adequate verbal and written handoff was measured in both periods (control and intervention), observing an increase in adherence in all the items after the application of the intervention (Fig. 2 & 3). The item “verbal handoff synthesis by the receiver” had the greatest adherence improvement, showing an increase from 1.3% to 87.6% in the intervention group, difference 86.3% (95% CI: 43.3 - 93.3), p = 0.0048]. The item written handoff illness severity compliance had the best improve [control 0.7% (0.0; 1.6) vs intervention 94.5% (87.0 - 97.7); difference 93.7% (95% CI: 78.4 - 98.3), p = 0.0048). The key elements of the verbal and written handoff are shown in table 3.

**Figure 2:**
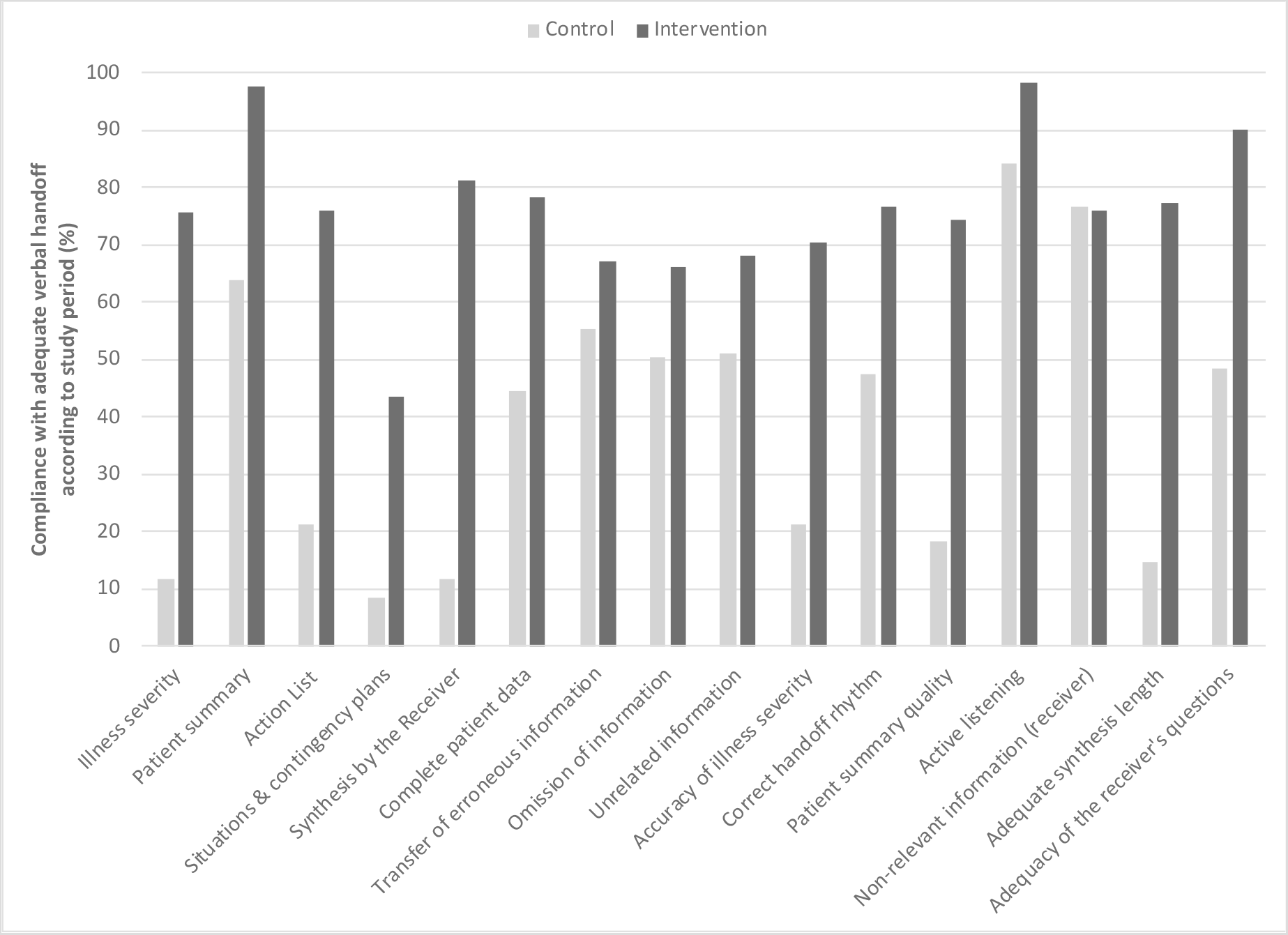
Percentage of verbal handoffs that included key data elements (all sites combined). Key elements evaluated for oral handoffs included an illness-severity assessment (unstable, watcher, stable), patient summary (defined as an oral handoff of at least three of the following: summary statement, events leading up to admission, hospital course, ongoing assessment, and active plans), action list (defined as a clearly articulated list of “to do” items or a statement of “nothing to do”), situations & contingency plans (defined as an indication of what to do if adverse contingencies occur, or an explicit indication that no adverse contingencies were anticipated), and synthesis by the receiver (defined as readback mostly performed with small correction required or readback fully performed without need for correction), complete patient data (defined as three identification including name), transfer of erroneous information (defined as information not belong to the patient or old fashioned), omission of information (defined as omission of more than one principal diagnoses or treatment), unrelated information (defined as information not related to the patient being handoff), accuracy of illness severity (defined as illness-severity assessment of the handoff giver and the one registered in the patient medical record), correct handoff rhythm (defined as normal rhythm of spoken voice without long interruptions), patient summary quality (as compared to the patient medical record), active listening(evaluates the receiver’s attention), non-relevant information (delivered by the receiver in the synthesis), adequate synthesis length (defined as no longer than 1 minute), adequate of the receiver’s questions (defined as questions related to patient state as compared to medical record).

**Figure 3:**
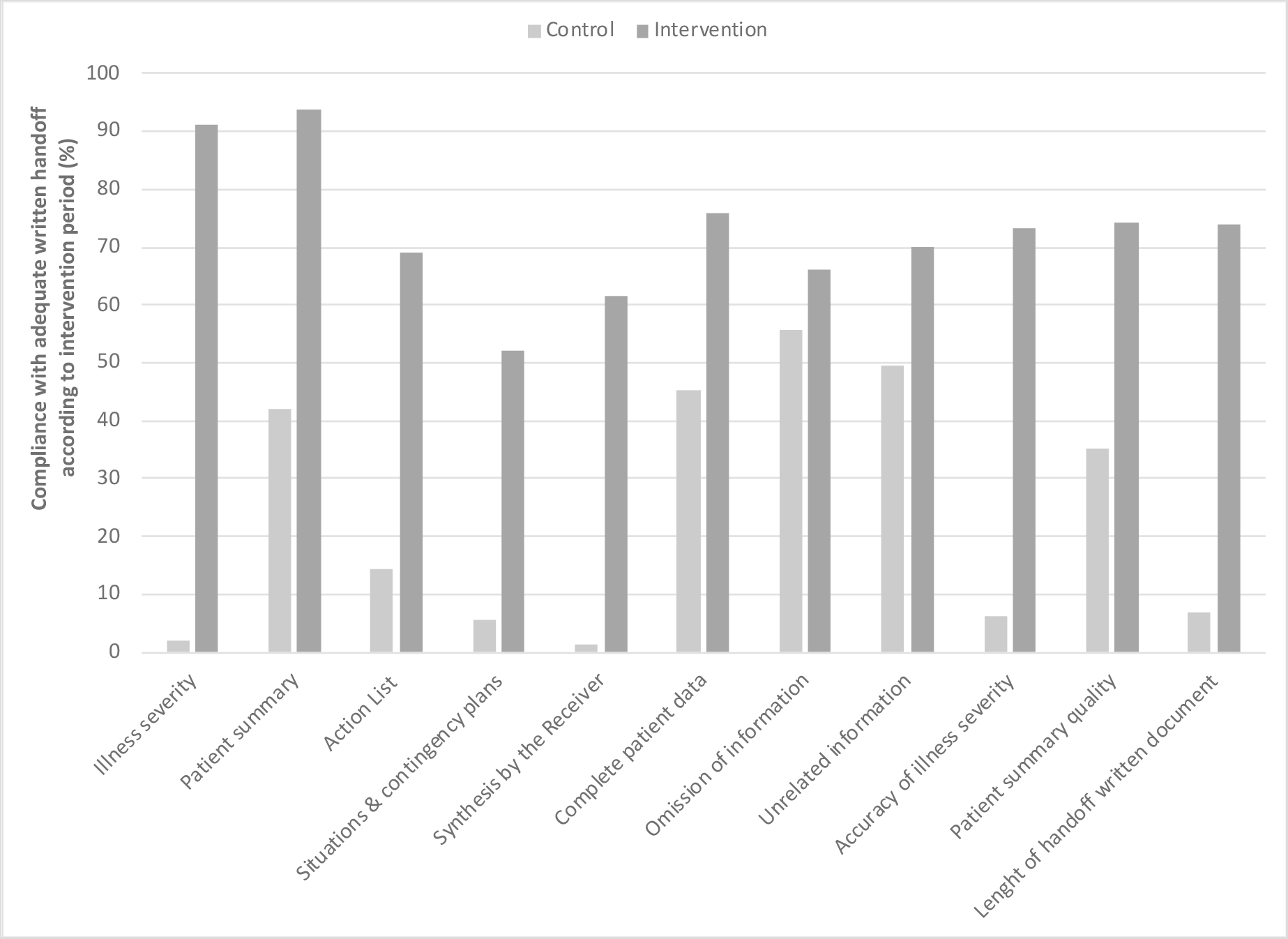
Percentage of written handoffs documents that included key data elements (all sites combined). Key elements evaluated for oral handoffs included an illness-severity assessment (unstable, watcher, stable), patient summary (defined as an oral handoff of at least three of the following: summary statement, events leading up to admission, hospital course, ongoing assessment, and active plans), action list (defined as a clearly articulated list of “to do” items or a statement of “nothing to do”), situations & contingency plans (defined as an indication of what to do if adverse contingencies occur, or an explicit indication that no adverse contingencies were anticipated), and synthesis by the receiver (defined as a remainder to do the synthesis in the document), complete patient data (defined as three identification including name), omission of information (defined as omission of more than one principal diagnoses or treatment), unrelated information (defined as information not related to the patient being handoff), accuracy of illness severity (defined as illness-severity assessment in the document and the one registered in the patient medical record), correct handoff rhythm (defined as normal rhythm of spoken voice without long interruptions), patient summary quality(as compared to the patient medical record), length of handoff written document.

Regarding the handoff duration, a longer time per patient was verified during the intervention period [control 5.96 minutes (4.69 - 7.23) vs intervention 7.29 minutes (5.77 - 8.81); difference 1.33 minutes (95% CI: 0.64 - 2.02), p = 0.0002], with no difference in the time spend in the handoff as a whole in both periods [35.7 minutes (29.6 - 41.8) vs intervention 34.7 minutes (26.5 - 42.1); difference 1.43 minutes (95% CI: 2.63 - 5.49), p = 0.4900].

### Patient safety culture survey

Eighty-two subjects answered in the control period and 87 in the intervention period, being the response rate was greater than 80% in each of the clusters. There was no difference in the positive answers between periods (Table 4).

**Table 4.**
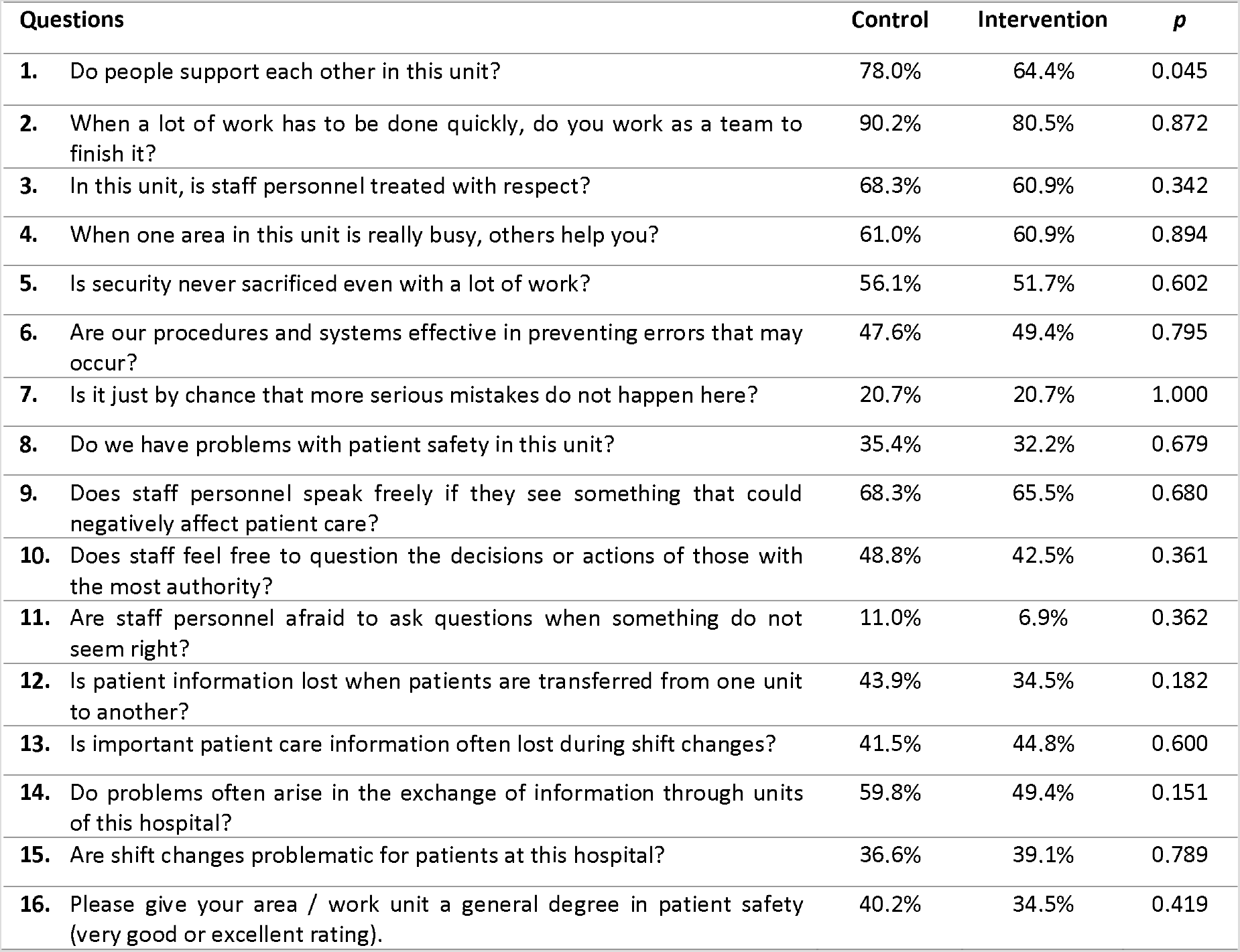
Results of the patient safety survey during both periods.

| Questions | Control | Intervention | *p* |
| --- | --- | --- | --- |
| 1. Do people support each other in this unit? | 78.0% | 64.4% | 0.045 |
| 2. When a lot of work has to be done quickly, do you work as a team to finish it? | 90.2% | 80.5% | 0.872 |
| 3. In this unit, is staff personnel treated with respect? | 68.3% | 60.9% | 0.342 |
| 4. When one area in this unit is really busy, others help you? | 61.0% | 60.9% | 0.894 |
| 5. Is security never sacrificed even with a lot of work? | 56.1% | 51.7% | 0.602 |
| 6. Are our procedures and systems effective in preventing errors that may occur? | 47.6% | 49.4% | 0.795 |
| 7. Is it just by chance that more serious mistakes do not happen here? | 20.7% | 20.7% | 1.000 |
| 8. Do we have problems with patient safety in this unit? | 35.4% | 32.2% | 0.679 |
| 9. Does staff personnel speak freely if they see something that could negatively affect patient care? | 68.3% | 65.5% | 0.680 |
| 10. Does staff feel free to question the decisions or actions of those with the most authority? | 48.8% | 42.5% | 0.361 |
| 11. Are staff personnel afraid to ask questions when something do not seem right? | 11.0% | 6.9% | 0.362 |
| 12. Is patient information lost when patients are transferred from one unit to another? | 43.9% | 34.5% | 0.182 |
| 13. Is important patient care information often lost during shift changes? | 41.5% | 44.8% | 0.600 |
| 14. Do problems often arise in the exchange of information through units of this hospital? | 59.8% | 49.4% | 0.151 |
| 15. Are shift changes problematic for patients at this hospital? | 36.6% | 39.1% | 0.789 |
| 16. Please give your area / work unit a general degree in patient safety (very good or excellent rating). | 40.2% | 34.5% | 0.419 |

## DISCUSSION

### Summary of findings

In this randomised stepped wedge trial in PICUs of Argentina, we assessed the effect of a standardized handoff intervention to reduce the frequency of AE and increase the quality of handoffs. The intervention resulted in an overall improvement in the quality of verbal and written handoffs. However, the intervention did not significantly affect the frequency of adverse events. Improvements were observed in all the items considered needed for a handoff, but it was greater in the key elements, like illness severity and synthesis by the receiver. Patient summary ranked positively in both periods. The majority of the handoff quality items were far below 50% compliance before the intervention, and some of them reached more than 90% of adherence after the I-PASS bundle implementation. The intervention was deployed in similar ways in all the sites.

### Strengths and limitations

This study had several strengths. We used a rigorous experimental design and achieved similar groups by using randomisation, which allowed the intervention to be evaluated effectively. The selected intervention components were previously documented as effective and were tailored according to formative research. Finally, to our knowledge this is the first trial that evaluates the implementation of I-PASS bundle and also the use of a trigger tool in paediatric patients.

Nonetheless, the study had some limitations. It is already known that direct observation moved handoffs from ‘backstage’ to ‘front-stage’, and residents performed handoffs differently from their usual practice when they were observed (23). Secondly, the intervention was evaluated immediately after its implementation and in some PICUs it was implemented only for four months. Thirdly, the periods of the year did not coincide exactly in each cluster, and the passage of time in improving handoffs cannot be ruled out. Fourthly, the cluster size reached were lower than calculated, due to a drop in the number of admissions in participating PICUs between November 2018 and January 2019. Finally, the study was only carried out in PICUs in the public subsector of one Latin American middle-income country, which prevents us from extrapolating the results to other populations.

### Interpretation

In this trial, the intervention did not significantly affect the total number of AE, the number of preventable AE, and the severity and categories of AE. Adverse Event rates are the product of numerous interacting institution structures and processes; and it is possible that variation in the ascertainment of error data or other unmeasured factors, were responsible for the lack of improvement in AE rates. We also found substantial inter-institutional variation in AE rates.

Regarding direct observation of health care providers, the majority of the observed staff personnel were physicians with many years of handoff experience, and we believe they were hardly influenced by being observed. Anyway, direct observation placed a spotlight on handoffs as a clinical skill, reinforcing the importance of doing it well.

Regarding the time used to carry out the handoff, no differences were observed with respect to the total time, and we observed a greater time spent per patient in the intervention stage. This difference could be due to the use of a new tool. Although a differential use of time towards other activities during the handoff cannot be ruled out, since the total time did not change in both periods. All the participating PICUs had paediatric residents or paediatric intensive care physicians in training, so the time of handoff was also used for teaching activities. This teaching activities often served to share mental model by providing the rationale for proposed management, which is an important feature of quality handoff (24,25). It has been suggested the I-PASS potential to reinforce an institutional culture that embraces interactive questioning and teaching opportunities with the goal of fostering shared understanding and optimizing patient care(26).

The AHRQ survey has been widely used in Spanish-speaking hospitals. None of the participating hospitals had previously used it in their PICUs. No changes were observed in the way physicians perceived patient safety related to the communication dimension, before and after implementing the intervention. One explanation may be the high baselines rates of patient safety and communication perception observed. The only one question with statistically significant difference was the one referred to mutual support. This item was not included in the teamwork training. The new situational awareness gained with the I-PASS bundle implementation could raise the necessity of more complete teamwork training.

### Comparison with previous literature

There is a lack of robust evidence on best handoff practices, and current knowledge on the nature of handoff failures during inter shift transfers is scanty. Starmer et al. observed that after the implementation of the I-PASS, a similar improvement was observed in the compliance of the items related to a quality handoff, while no differences were observed in the load of resident’s work. In this study, no changes were observed in the time used to transfer patients(7), and not all the sites reduced the AEs at the same level; also the compliance with the handoff key elements was heterogeneous. They observed a 23% relative reduction in medical errors and a 30% reduction in preventable AE. Decreased errors related to diagnosis, medical history and physical examination were observed. While they did not observe changes regarding errors related to medication, procedures or hospital infections (10). The AE report was based on direct observation or voluntary report. In our series, AEs were identified from MR, with a tool that, through pre-established triggers, can lead to AE, and also AE identified in the MR without triggers.

Shet et al. demonstrated that a process transfer supported by I-PASS was associated with better efficiency and culture of handoff safety(27). Coffey et al. in a study that showed experiences of residents with the implementation of the I-PASS package, they promoted the presence of other important active ingredients in this complex intervention, such as the automatic import of patient data in the electronic transfer document, improvements in the environment transfer, in teamwork and communication skills (12). They also noted that strict adherence may not be necessary to achieve the desired results. Like others, in our study, synthesis or re-reading was the most challenging feature of I-PASS for clinicians (10,13). The average compliance of the intervention items was similar to that observed in the different centres at the beginning of the implementation of the I-PASS program; a significant difference was also shown after the implementation of the quality improvement (14,15).

In a report of AE in hospital wards of 16 teaching and non-teaching hospitals it was also not possible to observe an improvement in the number of AE over time. The AE rate was higher in academic hospitals (26.2 AE per 1000 patient days [95% CI: 23.7–29.0]) (16). We recruited only teaching hospitals and to our knowledge this is the first study to use the GAPPS tool exclusively in PICUs, and the first one using the tools in Spanish. Although the number of AE was higher than that described by Stockwell et al. this study included patients with less severity than those evaluated in our study.

### Conclusions

In conclusion we observed an improvement in the quality of handoffs after the implementation of a standardized handoff intervention. No differences were observed in the AE after the use of I-PASS, nor in the perception of improvement in communication.

Further research is needed to determine whether this intervention could reduce AE either by different implementation models for a longer duration or using different outcomes measures.

## Data Availability

https://osf.io/gn4qu/?view_only=4472676a10a541e0be2f87f0971e7064

## Conflict of Interest

None to declare.

## Funding

Salud Investiga “Dr. Abraam Sonis” grant, category Multicentric Study, funded by National Ministry of Health, through the Directorate of Research for Health, Argentine.

The funders of the study had no role in study design, data collection, data analysis, data interpretation, or writing of the report.

## Acknowledgments

The author would like to thank specially Amy Starmer for her assistance with the adaptation of the I-PASS bundle and Christopher Landrigan for his assistance with the GAPPS tool use. Also, we would like to thank to the staff of the Hospital General de Niños Pedro de Elizalde, Hospital General de Niños Ricardo Gutierrez, Units 44 and 45 of the Hospital Nacional de Pediatría “J. P. Garrahan”, Hospital de Niños de San Justo and Hospital Materno Infantil “Héctor Quintana”.

Primary data that supports these findings is available at Open Science Framework. Final D.O.I. to be released on publication.

## Clinical Trials Registration Number

NCT03924570

## Ethical approvals

the study was reviewed and approved by all the 5 hospitals Institutional Review Boards.

## Notes

### Competing Interest Statement

Dr. Jorro-Baron, Dr. Landry, Dr. Penazzi, Dr. Rodriguez, G, Dr. Demaio, Dr. Burgos-Pratx and Dr. Suarez-Anzorena report grants from Ministry of Health of the Nation, through the Directorate of Research for Health, Argentine, during the conduct of the study.

### Clinical Trial

NCT03924570

### Clinical Protocols

https://clinicaltrials.gov/ct2/show/study/NCT03924570

### Author Declarations

Hospital General de Ninos Pedro de Elizalde, Comite de Etica en Investigacion 05/06/2018 Hospital General de Ninos Ricardo Gutierrez, Comite de Etica en Investigacion CEI Nro 18.18

